# Beyond Doppler: Scalable AI Detection of LVOT Obstruction in HCM

**DOI:** 10.64898/2026.04.17.26351151

**Authors:** Owen R. Crystal, Juan M. Farina, Isabel G. Scalia, Chadi Ayoub, Hyung Bok Park, Kyung An Kim, Reza Arsanjani, Steven J. Lester, Imon Banerjee

**Affiliations:** Department of Cardiovascular Medicine, Mayo Clinic, Phoenix, Arizona, USA; Division of Cardiology, Department of Internal Medicine, International St. Mary’s Hospital, Catholic Kwandong University College of Medicine, Incheon, South Korea; Department of Cardiology, Incheon St. Mary’s Hospital, The Catholic University of Korea, Seoul, South Korea; Department of Radiology, Mayo Clinic, Phoenix, Arizona, USA

**Keywords:** Left ventricular outflow tract gradient, hypertrophic cardiomyopathy, echocardiography, deep learning, multi-view fusion, artificial intelligence

## Abstract

**Background:** Accurate assessment of left ventricular outflow tract (LVOT) gradients is critical for hypertrophic cardiomyopathy (HCM) management, yet Doppler-based measurements are technically demanding and require expertise.

**Objective:** To develop a multi-view deep learning model capable of classifying LVOT obstruction (> 20mmHg) using routine 2D echocardiographic windows without reliance on Doppler imaging.

**Methods:** We trained and externally validated a cross-attention-based video-to-video fusion framework that integrated EchoPrime-derived video representations from three standard transthoracic echocardiographic views to classify LVOT gradients.

**Results:** Training was performed on a derivation cohort (N = 1833) from a tertiary care system in the United States, with model performance evaluated on an internal held-out test set (N = 275) and a Korean external validation cohort (N = 46). Single-view baselines showed limited discrimination (external AUROCs 0.47–0.70). Conversely, domain-specific foundational model (EchoPrime) achieved superior single-view performance (AUROCs 0.75–0.80 internal; 0.79–0.83 external), highlighting the importance of echo-specific pretraining and temporal modeling. The proposed multi-view fusion further enhanced predictive performance, with the late fusion model reaching an AUROC of 0.84 on the external cohort with significant population-shift.

**Conclusions:** These results suggest LVOT physiology is encoded in routine 2D imaging and can be leveraged for clinically relevant gradient classification without Doppler input- proposed AI-guided strategy demonstrates substantial cost savings compared with the screen-all approach. By integrating complementary spatial-temporal information across multiple views, our approach generalizes robustly across populations and may enable real-time decision support, extend LVOT assessment to portable or resource-limited settings, and complement Doppler-based evaluation for longitudinal HCM management.

## Introduction

Hypertrophic cardiomyopathy (HCM) is a prevalent genetic cardiomyopathy with a heterogeneous clinical presentation and variable outcomes^1^. Approximately two-thirds of the patients with HCM have dynamic left ventricular outflow tract obstruction (LVOTO), a major determinant of symptoms, functional limitations, and prognosis. Defining the presence and severity of LVOTO plays a central role in therapeutic decision-making including the selection of invasive interventions and emerging targeted treatments^2–4^. Accurate quantification of the left ventricular outflow tract (LVOT) gradient is therefore essential in the routine evaluation and longitudinal management of patients with HCM^5^.

Transthoracic echocardiography (TTE) is the primary imaging modality for assessing LVOTO. Spectral Doppler-based LVOT gradient quantification requires multi-view acquisition to ensure parallel alignment of the continuous wave Doppler beam with the LVOT flow vector, underscoring its operator dependence^2,5–7^. As a result, gradient measurement and reproducibility of the assessment remains challenging, particularly in settings where access to highly skilled echocardiography operators is limited. Collectively, these limitations underscore the need for an accessible, and scalable multi-view echocardiography–based LVOT gradient classification framework.

Deep learning (DL) methods have demonstrated effectiveness in automating diagnostic image interpretation enabling more efficient and scalable workflows across medical imaging modalities^8–13^. Although DL has been successfully applied to numerous echocardiographic tasks, automated non-Doppler-based classification of LVOTO remains understudied. Development of a fully automated, multi-echocardiographic view, image-based method capable of predicting Doppler-derived LVOT gradients in patients with HCM would have profound clinical utility.

In this study, we sought to develop and validate a fusion DL model using multi-view non-Doppler TTE images to predict LVOT gradients above or below 20mmHg. By eliminating the need for Doppler-based images and manual measurements, this approach aims to provide a reproducible, scalable tool that will streamline LVOT assessment and improve accessibility in diverse clinical environments.

## Methods

### Cohort selection

Approval from Mayo Institutional Review Board was obtained. All TTE studies acquired between 1/1/2010 and 1/27/2023 across a multi-center tertiary care system (Mayo Clinic Rochester, Arizona, Florida and Health System) in the United States were retrospectively screened for the presence of HCM. Patients were identified through the electronic health record using relevant International Classification of Diseases (ICD) diagnostic codes for HCM. TTEs were randomly divided into train, validation, and test sets with a 70/15/15 percent patient-level split, respectively. An external validation dataset was obtained from International St. Mary’s Hospital (Seo-gu, Incheon, South Korea) comprising exclusively patients of Asian race, with all echocardiographic studies acquired using GE Ultrasound systems.

The presence and severity of LVOTO were quantified from a resting TTE by using spectral Doppler in accordance with current guidelines^1,14^. Color Doppler was used to identify the outflow jet and optimize alignment of the continuous wave Doppler beam as parallel as possible to the LVOT flow direction. LVOTO was considered significant if greater than 20 mmHg. A maximal instantaneous LVOT gradient cutoff of 20 mmHg was selected because it is practically and clinically applicable, aligning with the threshold validated in clinical trials to reflect stability of HCM obstruction in patients treated with myosin inhibitor therapy ^15^. We acknowledge that earlier literature commonly used a cutoff of 30 mmHg to define the presence of obstruction. The obtained gradients were recorded as continuous variables in mmHg. When provocative maneuvers were performed (either the strain phase of the Valsalva maneuver, squat to stand or amyl nitrite inhalation), the highest recorded LVOT gradient was used for analysis. After obtaining LVOT gradients as a continuous variable expressed in mmHg, it was converted into a binary variable using 20 mmHg as the cutoff point to determine the presence/absence of significant LVOTO. Images and gradients were obtained by expert cardiologists with board certification in echocardiography.

### Image preprocessing and view selection

All TTE studies underwent a standardized preprocessing pipeline prior to model training and inference. Using a pre-trained and previously validated deep learning model^16,17^, each echocardiogram video was automatically classified into one of the six standard transthoracic echocardiographic views: parasternal long axis (PLAX), parasternal short-axis at the papillary muscle level, parasternal short-axis at the mitral level, apical two-chamber, apical three chamber (AP3), and apical four-chamber (AP4). For this study, AP3, AP4, and PLAX were selected. AP3 and PLAX were selected as they provide clear views of the mitral valve and any systolic anterior motion that may occur with obstruction, as well as the LVOT. The AP4 view was included to complement these focused views by providing a global assessment of left ventricular geometry, septal thickness, and concurrent atrioventricular morphology. Only studies with a PLAX, AP4, or AP3 view classification and a predicted probability ≥ 0.80 were retained for downstream analysis. The threshold was determined using the internal validation dataset to favor high-confidence predictions and minimize view misclassification, thereby ensuring reliable and standardized view selection for subsequent modeling.

### Model architecture

We developed an end-to-end framework leveraging a pre-existing 3D foundation model backbone (EchoPrime^18^), a bi-directional attention-based multi-view fusion strategy to effectively capture complementary information across multiple echocardiogram views, and a late fusion technique to incorporate all three echo imaging views in a meaningful manner (Figure 1).

**Figure 1.**
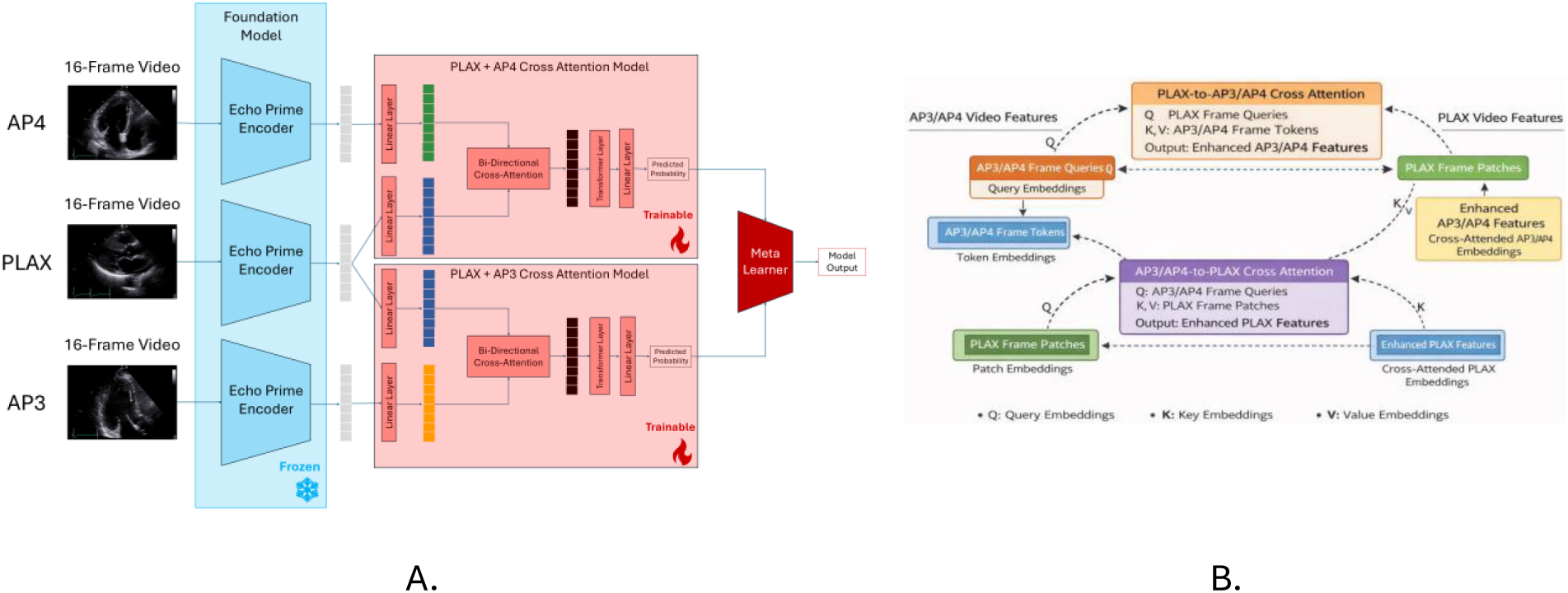
Architectural diagram - linear probing of foundational model with the attention-based multi-view fusion for leveraging complimentary strength of multiple echocardiographic views in a study. Predictions from the two multi-view fusion blocks are used as input features into a final meta learner. - (A) end-to-end framework diagram, (B) bi-directional cross-attention inner working scheme.

#### Video encoding using foundational model

For encoding, we leveraged EchoPrime, a recently developed 3D echocardiography foundation model^18^. EchoPrime is a video-based vision– language model trained on 12,124,168 echocardiography videos paired with clinical text reports from 275,442 studies spanning 108,913 patients at Cedars-Sinai Medical Center and consists of a video encoder. We selected 32 contiguous frames from each TTE video, and to approximate temporal down sampling while preserving motion dynamics, every other frame from this segment was selected, yielding a 16-frame sequence that served as the model input. The echocardiographic frames were cropped and masked following EchoPrime^18^, ensuring consistent spatial focus across samples. We adopted the EchoPrime video encoder as a frozen feature extractor, generating fixed embeddings (N = 512) for each echocardiographic view. Specifically, AP4, AP3, and PLAX views were encoded independently to obtain view-specific video-level feature representations.

#### Attention-based multi-view fusion

To capture complementary information across multi-view echocardiographic studies, we developed an innovative bidirectional view cross-attention and evaluated it against multiple standard fusion strategies trained using feature-level concatenation, and decision-level meta-classification. In the bidirectional cross-attention framework, embeddings from each echocardiographic view were treated as both queries and keys/values for the other views, enabling mutual information exchange across views (Figure 1B). Clinically, this design mirrors expert echocardiographic interpretation, where the parasternal long-axis (PLAX) view often serves as the primary reference for mitral valve evaluation, while apical views (AP3 and AP4) provide complementary functional and hemodynamic information. Given the central role of the PLAX view, cross-attention was implemented using the PLAX view embedding as the anchor, enabling information exchange between PLAX–AP3 and PLAX–AP4 representations. In this framework, the PLAX embedding attends to features from the apical views and, conversely, the apical embeddings attend to PLAX features, allowing mutual refinement of representations across views. This configuration enables the model to dynamically weight and integrate complementary spatial and temporal information from multiple echocardiographic perspectives, reinforcing clinically salient patterns while suppressing redundant or low-quality signals.

The attended representations were subsequently aggregated and propagated to downstream classification layers, enhancing cross-view contextual understanding and improving patient-level predictions prior to final inference.

#### Decision-level late fusion

To further leverage complementary strengths across the two attention-based models, we implemented a decision-level meta-learner that uses the predicted outputs (continuous probabilities) from the PLAX+AP3 and PLAX+AP4 models. This approach enables the meta-learner to learn the optimal weight for combining predictions from each view-pair, allowing adaptive prioritization of the most informative representation for each patient.

The training was performed on 4 core NVIDIA RTX A6000 using the Adam optimizer with a learning rate of 1×10⁻⁴, weight decay of 0.001, batch size of 32, using weighted binary cross entropy loss for 100 epochs. Early stopping with a patience of 10 epochs based on the validation loss was used to prevent overfitting. The hyperparameters of the logistic regression meta-classifiers were optimized using a grid search.

### Statistical Evaluation

We benchmarked the performance of the proposed framework against multiple baseline models. These included a standard 2D convolutional neural network based on DenseNet121^19^ initialized with ImageNet weights^20^, as well as linear probing approaches using a frozen 2D vision transformer foundation model (MedCLIP) as a feature extractor^21^. For 2D-based models, frame-level predictions were generated at inference and averaged across all frames within a study to obtain study-level predictions. As a 3D convolutional baseline, we employed the ResNet(2+1)D-18 (R(2+1)D-18) architecture^22^, which uses factorized 2D spatial and 1D temporal convolutions to jointly capture structural and temporal dynamics in echocardiographic videos. This model was initialized with Kinetics-400 weights^23^. For comparative evaluation of multi-view fusion strategies, we additionally benchmarked against a logistic regression meta-classifier that was trained using probabilistic decision from all three single-view EchoPrime linear probe models.

All models were trained/fine-tuned using the same split and evaluated on the held-out internal test set and an external validation set. The performance was analyzed based on Area Under the Receiver Operating Curve (AUROC). Model calibration was assessed using calibration curves and the Brier score to evaluate agreement between predicted probabilities and observed outcomes. A 95% confidence interval was calculated for each metric using bootstrapping with 100 iterations. Two-sided t-tests were performed to compare baseline performance metrics to the proposed model.

## Results

### Quantitative Results

In total, 1,833 patients were included in the internal cohort (1387, 75.6% with a LVOT gradient > 20 mmHg) and 46 in the external cohort (12, 26.1% with an LVOT gradient > 20 mmHg). Table 1 describes the characteristics of the internal and external datasets. Table 2 summarizes the performance of the baseline and proposed models on both cohorts. Figure 2 shows the ROC curves, sensitivity-specificity versus thresholds plots, and calibration curves for the proposed attention-based models. Supplementary Figure 1 shows the ROC curves for the baseline models.

**Figure 2.**
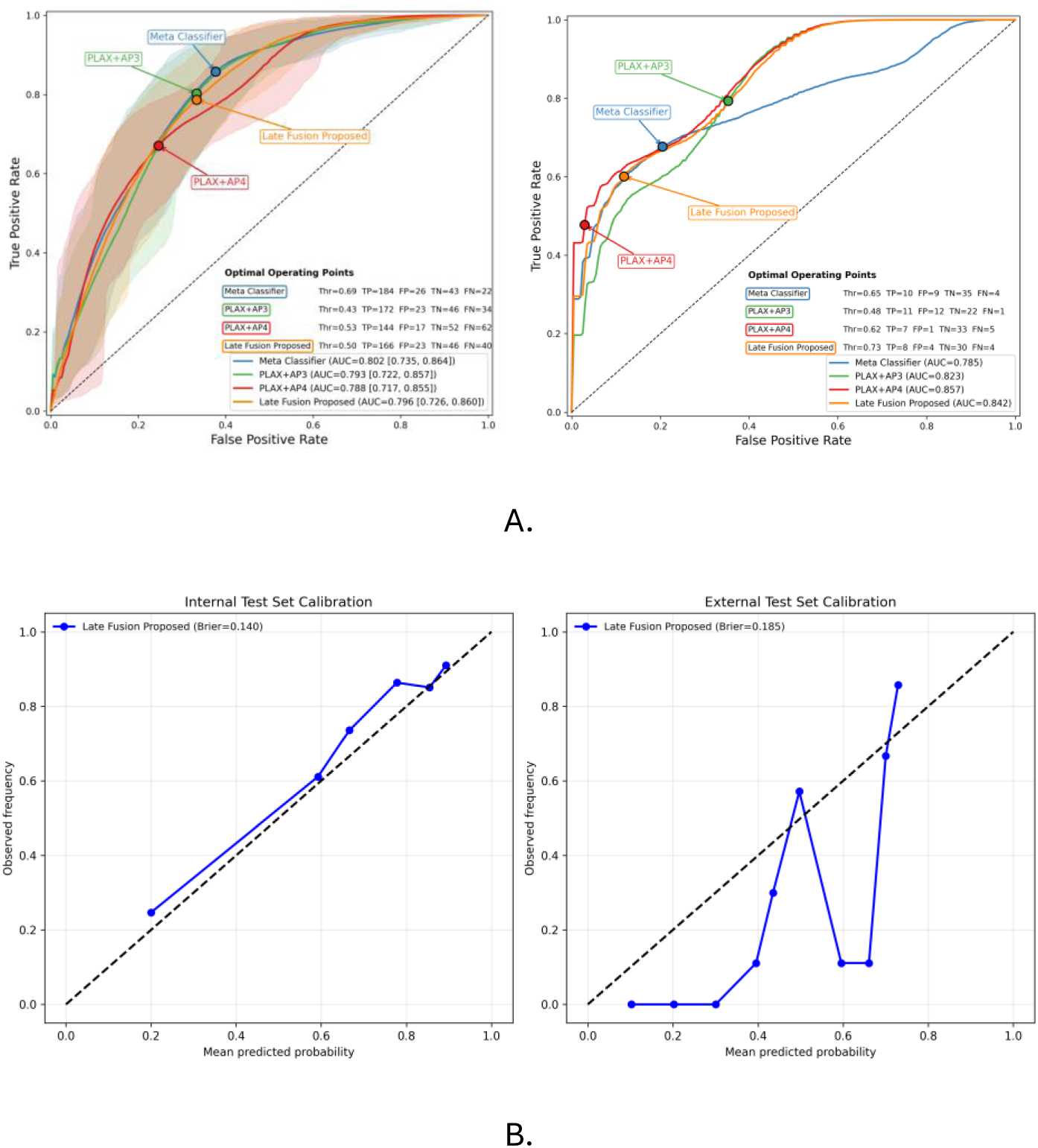
Quantitative performance - (A) ROC curves for the fusion models for the internal (left) and external (right) test set. Optimal operating points and the corresponding number of true positives (TP), false positives (FP), true negatives (TN), and false negatives (FN) are listed for each model; (B) Calibration curves for proposed bi-directional attention-based models on the internal and external cohorts.

**Table 1.**
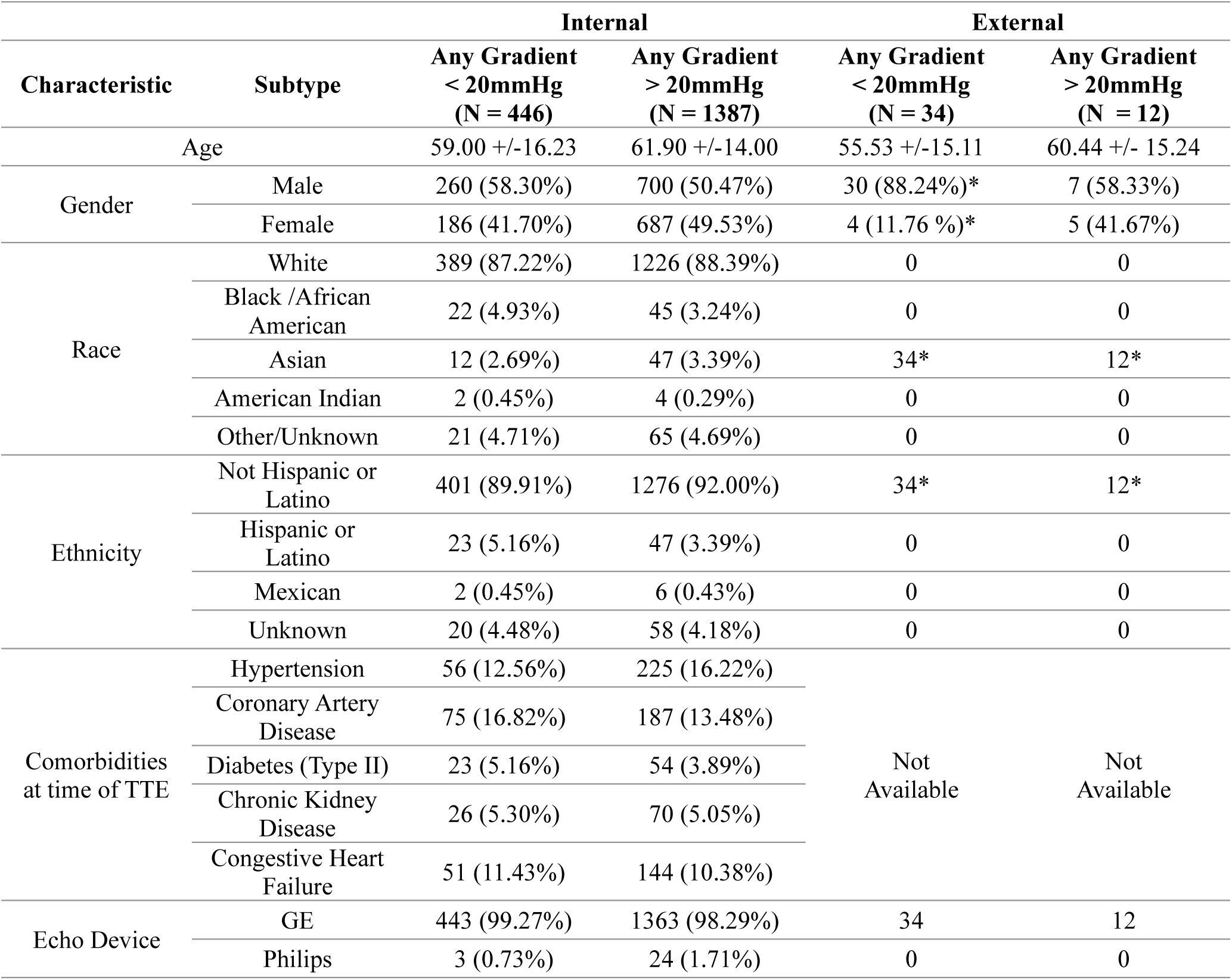
Cohort characteristics of internal and external datasets.

**Table 2:** Quantitative performance comparison between the proposed framework and 2D/3D CNN baselines, and foundation models.

| Model Group | Architecture | Model | Internal (N = 275) |  |  | External (N = 46) |  |  |
| --- | --- | --- | --- | --- | --- | --- | --- | --- |
|  |  |  | AUROC | Sens | Spec. | AUROC | Sens. | Spec. |
| CNN | DenseNet121<sup>19</sup> | AP4 DenseNet121 | 0.70 [0.62, 0.78]* | 0.78 [0.72, 0.83] | 0.59 [0.52, 0.65] | 0.63* | 0.58 | 0.76 |
|  |  | AP3 DenseNet121 | 0.69 [0.61, 0.77]* | 0.84 [0.77, 0.90] | 0.49 [0.41, 0.56] | 0.60* | 0.67 | 0.62 |
|  |  | PLAX DenseNet121 | 0.71 [0.62, 0.79]* | 0.87 [0.82, 0.93] | 0.52 [0.46, 0.58] | 0.53* | 0.75 | 0.50 |
|  | ResNet18-3D<sup>22</sup> | AP4 R(2+1)D-18 | 0.55 [0.44, 0.65]* | 0.37 [0.31, 0.44] | 0.74 [0.69, 0.79] | 0.67* | 0.75 | 0.71 |
|  |  | AP3 R(2+1)D-18 | 0.55 [0.45, 0.64]* | 0.26 [0.20, 0.32] | 0.87 [0.81, 0.92] | 0.70* | 1.00 | 0.38 |
|  |  | PLAX R(2+1)D-18 | 0.58 [0.47, 0.70]* | 0.64 [0.58, 0.69] | 0.59 [0.52, 0.65] | 0.53* | 0.83 | 0.35 |
| Foundation | MedCLIP<sup>21</sup> | AP4 MedCLIP | 0.63 [0.56, 0.71]* | 0.54 [0.47, 0.61] | 0.72 [0.66, 0.78] | 0.53* | 0.92 | 0.15 |
|  |  | AP3 MedCLIP | 0.63 [0.55, 0.70]* | 0.59 [0.53, 0.65] | 0.67 [0.61, 0.72] | 0.56* | 1.00 | 0.12 |
|  |  | PLAX MedCLIP | 0.65 [0.57, 0.73]* | 0.70 [0.62, 0.77] | 0.58 [0.51, 0.66] | 0.47* | 0.08 | 1.00 |
|  | EchoPrime<sup>18</sup> | AP4 Echo Prime | 0.75 [0.68, 0.82]* | 0.69 [0.63, 0.75] | 0.71 [0.66, 0.77] | 0.80* | 0.58 | 0.91 |
|  |  | AP3 Echo Prime | 0.76 [0.69, 0.83]* | 0.85 [0.78, 0.91] | 0.62 [0.56, 0.69] | 0.83 | 0.83 | 0.76 |
|  |  | PLAX Echo Prime | **0.80 [0.74, 0.86]** | **0.89 [0.83, 0.94]** | 0.59 [0.52, 0.65] | 0.79* | 0.58 | 0.91 |
| Fusion |  | 3-View MC | **0.80 [0.73, 0.86]** | **0.89 [0.83, 0.94]** | 0.62 [0.56, 0.68] | 0.78* | 0.71 | 0.90 |
|  |  | PLAX + AP4 X-Attn | 0.79 [0.72, 0.86] | 0.70 [0.64, 0.76] | 0.75 [0.69, 0.81] | **0.86** | 0.58 | 0.97 |
|  |  | PLAX + AP3 X-Attn | 0.79 [0.72, 0.86] | 0.83 [0.77, 0.88] | 0.67 [0.62, 0.74] | 0.82 | 0.92 | 0.65 |
|  |  | Late Fusion Proposed | **0.80 [0.73, 0.86]** | 0.81 [0.75, 0.87] | 0.67 [0.62, 0.74] | 0.84 | 0.67 | 0.88 |

*Note.* Performance metrics are reported with 95% confidence intervals (in []), estimated using bootstrap resampling. Asterisks (\*) indicate statistically significant differences ( $p < 0.05$ ) relative to baseline models. Best-performing results for each cohort are shown in **bold**.

We first evaluated frame-based 2D models and single-view video models to establish baselines, which were then compared to the proposed multi-view fusion approaches. Across all echocardiographic views, the 2D DenseNet121 models achieved moderate internal performance, with AUROCs ranging from 0.69-0.71. Linear probing using the MedCLIP foundational vision encoder performed slightly worse (AUROCs 0.63-0.65), despite its extensive medical data pretraining. The R(2+1)D-18 model yielded the lowest overall performance (AUROCs 0.55-0.58), indicating that the available dataset (∼2,000 samples) was insufficient to train a convolutional neural network (CNN) despite initializing the model with Kinetics-400 weights.

In contrast, the single-view EchoPrime foundation models with simple linear probing significantly outperformed all baselines, achieving AUROCs of 0.75 (AP4), 0.76 (AP3), and 0.80 (PLAX) on the internal dataset. These results highlight the importance of large-scale, echo-specific pretraining and explicit temporal modeling of relevant hemodynamic features for LVOTO identification.

Performance difference between the EchoPrime-based models and the baselines widened significantly on the external dataset. DenseNet121 models demonstrated reduced discrimination (AUROCs 0.64-0.70), and MedCLIP linear probes also showed further decline in performance (0.47, 0.56). R(2+1)D-18 models remained limited with AUROCs of 0.53-0.70, demonstrating generalization challenges for frame-based architectures and 3D CNNs with limited samples.

EchoPrime linear probes maintained their strong performance with AUROCs of 0.80 (AP4), 0.83 (AP3), and 0.79 (PLAX) showing a strong ability to generalize external data.

Multi-view fusion models were developed to leverage complementary anatomical and temporal information across multi-view echocardiographic images. When testing the internal held-out test set, all fusion strategies matched or exceeded the best single-view performance. The 3-view logistic regression meta-classifier achieved an AUROC of 0.80 [0.73, 0.86], while the cross-attention models (PLAX+AP4 and PLAX+AP3) both reached an AUROC of 0.79. The proposed late fusion model demonstrated comparable internal performance (AUROC = 0.80) while improving the balance between false positives and false negatives. Specifically, the PLAX+AP4 cross-attention model reduced false positives but exhibited a higher false negative rate, whereas the PLAX+AP3 model showed the opposite pattern, with fewer false negatives at the expense of increased false positives. By integrating predictions from both view-pair models, the late fusion approach effectively balanced these complementary error profiles. The advantages of multi-view integration were evident on the external dataset as the PLAX+AP4 cross-attention model achieved the highest AUROC of 0.86. The proposed late fusion model achieved strong performance (AUROC = 0.84) while maintaining a more favorable tradeoff between sensitivity and specificity.

Collectively, these results demonstrate that LVOTO classification is feasible using non-Doppler echocardiographic videos. While 2D and 3D CNNs exhibit limited generalization, EchoPrime-based models provide strong single-view performance. The proposed fusion model further improved performance as they captured valuable complementary spatial-temporal information across views while preserving the generalization capability of the domain specific EchoPrime foundation model.

### Cost-benefit analysis

Figure 3 illustrates a decision tree comparing the proposed fusion model with a “screen-all” strategy in which all 1,000 patients undergo comprehensive rest and provocative TTE (CRPT) for evaluation of elevated LVOT gradients. Notably, the decision tree does not account for real-world constraints of the conventional approach, including the need for specialized centers with limited availability and associated delays in access. All costs were estimated based on clinical practice at Mayo Clinic (United States) and may not generalize as is to other healthcare settings and TP, FP, and FN is calculated based on current reported performance on the external validation dataset. In the AI-guided strategy, patients are first screened for hypertrophic cardiomyopathy (HCM) using a previously validated AI model^17^, and those classified as positive are subsequently evaluated using the proposed GradientAI model for LVOT gradient assessment. This sequential approach reduces the number of patients requiring advanced testing. The AI-guided strategy demonstrates substantial cost savings (∼3-fold reduction) compared with the screen-all approach ($1,104,000 vs. $3,000,000), while maintaining clinical performance, supporting its potential to improve efficiency and resource utilization. An additional version of this decision tree analysis using Medicare/Medicaid-based cost estimates is shown in Supplementary Figure 2.

**Figure 3.**
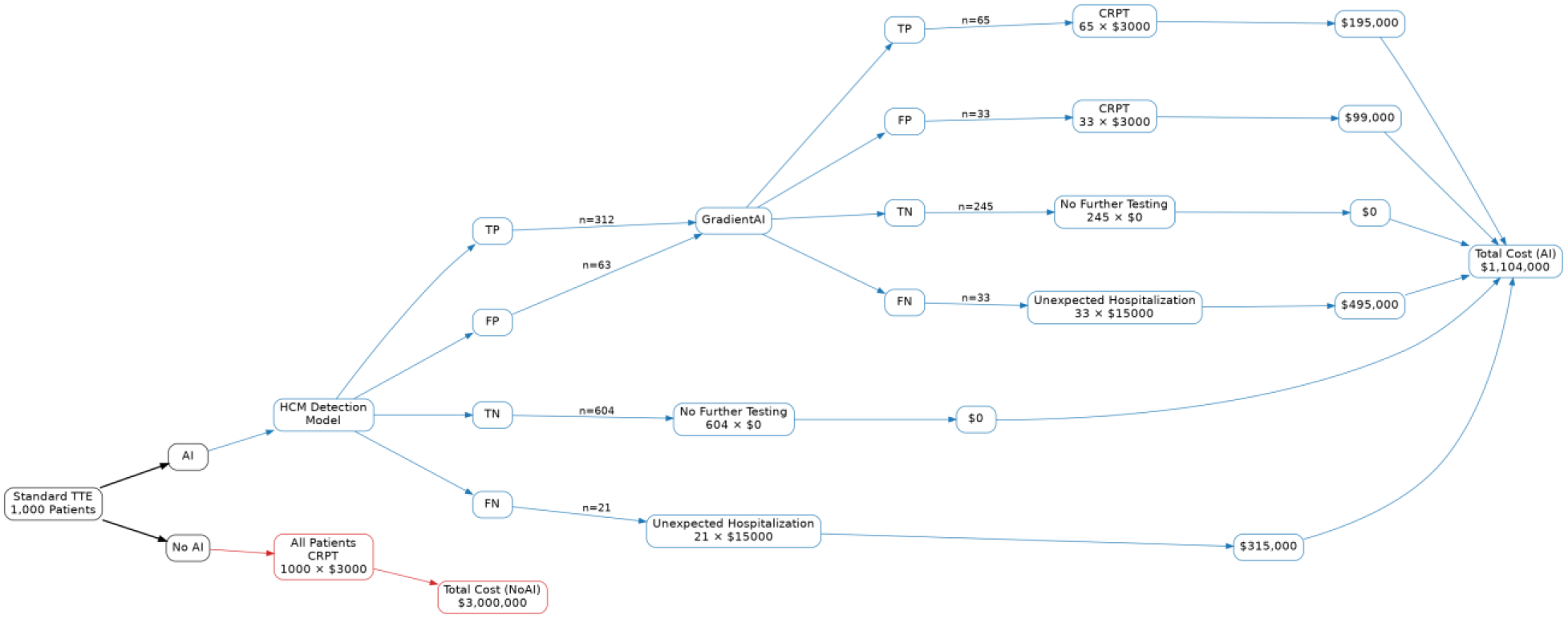
Decision tree analysis evaluating the financial impact of a two-stage AI-guided screening strategy for hypertrophic cardiomyopathy (HCM) triage and screening for elevated LVOT gradients. Outcomes per 1,000 patients undergoing standard transthoracic echocardiography (TTE) were estimated based on prevalence in the external test set. In the AI-guided strategy, patients first undergo HCM detection^17^, after which those classified as positive are referred to the proposed model (GradientAI) for LVOT gradient assessment. Patients are stratified into true positive (TP), false positive (FP), true negative (TN), and false negative (FN) groups across both stages, with associated costs assigned to each outcome. Positives cases received a comprehensive rest and provocative TTE (CRPT). In the “No AI” strategy, all patients undergo full echocardiogram with provocable maneuvers when needed, resulting in no missed cases but increased downstream testing. Total costs for each strategy are shown.

### Qualitative Analysis of Errors

Explainability is challenging for fusion models because the model’s reasoning is distributed across modalities, context-dependent, and nonlinearly entangled. Thus, we generated saliency maps using an occlusion sensitivity approach where in each input video (multi-view), localized regions were masked to quantify their contribution to the model’s output^24^. A sliding window of size 4 x 16 x 16 (frames x height x width) with a stride of 4 x 16 x 16 was applied to the input video to ensure non-overlapping patches were masked. These occlusion maps were normalized and represented as heatmaps overlaid onto the original echocardiographic video to provide visual interpretation.

Occlusion maps for the proposed attention-based models are shown in Figure 4. A threshold was applied to ensure that only attention values greater than 0.60 were shown in the overlay. A true positive and false positive example was provided along with their corresponding LVOT gradients and predicted probabilities for each echocardiographic view. Across the true positive cases, the model consistently focused on anatomically and physiologically relevant areas at end-systole including the LVOT, interventricular septum, and mitral valve when correctly identifying LVOTO. This localization pattern suggests that the model relies on meaningful physiological and structural cues when properly detecting LVOTO.

**Figure 4.**
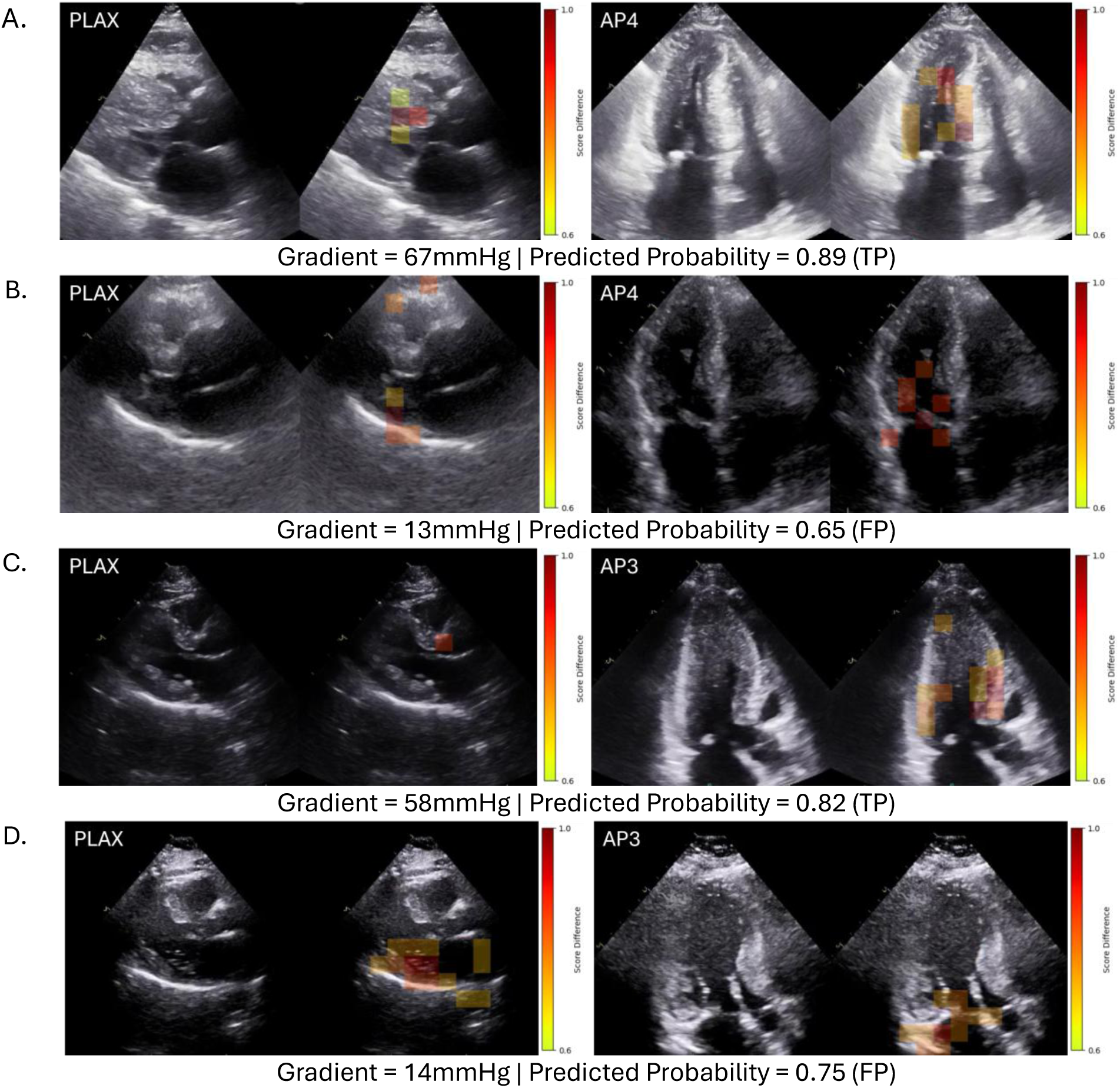
Single-frame occlusion map visualizations. True positive (A) and false positive (B) case using PLAX+AP4 model; True positive (C) and false positive (D) case using PLAX+AP3 model. The overlaid heatmaps demonstrate that the model’s attention is highly concentrated on physiologically relevant anatomical structures, specifically the interventricular septum, the mitral valve apparatus, and the LVOT itself.

Although the model frequently attended the same core anatomical regions on the false positive cases, the focused areas appeared more diffuse and general in comparison to the true positives. Additionally, the false positives shown in Figure 4 had peak LVOT gradients of 13, and 14mmHg, values close to the 20mmHg threshold. Misclassification of these cases may reflect physiological ambiguity associated with LVOTO rather than inherent model error. This claim is further supported by the supplemental case (see Supplementary Figure 3), where the Doppler showed moderate increase in LVOT gradient (peak velocity ∼1.6m/s) but did not meet the threshold for LVOTO despite showing signs of early or borderline obstruction.

## Discussion

We developed and externally validated a DL framework capable of classifying LVOT gradients in patients with HCM using only 2D, non-Doppler videos from standard TTE views. Several key findings emerged. First, integrating information across standard TTE views improved predictive performance, specifically on the external dataset, with cross-attention fusion of PLAX and apical views achieving the highest discrimination for elevated LVOT gradients. Second, domain-specific EchoPrime, an echocardiography foundation model pretrained on large-scale video data, consistently outperformed all conventional convolutional neural network baselines across views and a 2D generic foundation model (MedCLIP). Third, the performance of the proposed fusion model in the external cohort with significant population drift (∼3% Asian in training; 100% Asian in external) suggests strong generalizability beyond the development environment, with joint fusion models demonstrating superior generalizability compared with the late fusion approach.

These results demonstrate that meaningful structural and dynamic cues related to LVOT physiology are embedded within routine imaging and can be leveraged for clinically relevant gradient classification using DL architectures. Accurate LVOT gradient assessment is central to HCM evaluation and guides symptom interpretation, risk stratification, and therapeutic decision-making, including pharmacologic therapy, and interventional procedures^1,25,26^. Conventional Doppler assessment requires careful alignment of continuous-wave signals across multiple windows^5^. These steps are technically demanding, operator-dependent, and time-intensive, and may be difficult to implement in settings where comprehensive Doppler assessment is not readily available^6,7^.

Early AI-based efforts to quantify LVOTO in patients with HCM have predominantly relied on clinical variables and echocardiographic metrics, to train traditional machine learning models for predicting gradients^27,28^. More recent DL approaches have shown that severe LVOTO can be detected from single-view B-mode videos (PLAX), achieving strong performance in internal and external cohorts^29^. These studies provided initial evidence that LVOT physiology may be partially inferred from 2D motion patterns even without Doppler input. Previous work has also examined LVOTO classification using cardiac magnetic resonance imaging^30^, but these methods depend on manually engineered features, which are time-consuming to generate and inherently prone to inter-observer variability.

Our work expands this field in three important ways. First, by incorporating multiple echocardiographic views, we demonstrate that fusing complementary spatial-temporal information helped us to mimic the expert-read and ultimately yields higher performance than any single view alone. This mirrors clinical practice, in which PLAX, AP3, and AP4 views are collectively interpreted to understand the interplay between septal morphology, mitral valve motion, and LVOT geometry. Second, unlike prior models optimized solely for severe obstruction detection (≥50 mmHg), our approach is trained to classify gradients above or below 20 mmHg, showing that a broader range of LVOT loading conditions can be identified from 2D imaging. Additionally, the use of this cutoff point could have practical implications to define and adjust individualized medications and dose adjustment^15^. Third, domain-specific EchoPrime’s consistently outperformed other architectures (even medical imaging foundational model – MedCLIP), which highlights the value of large-scale pretraining and explicit temporal modeling. EchoPrime is pretrained on large-scale data drawn directly from the target domain, enabling them to learn representations that capture domain-relevant structure, texture, temporal dynamics, and context. In contrast, MedCLIP pretrained on gray scale non-echocardiographic medical images (primarily chest X-ray) often fail to encode subtle, domain-specific patterns critical for downstream tasks (e.g., cardiac motion patterns or lesion morphology). These elements appear essential for stable generalization beyond the development dataset.

The observed performance hierarchy offers insights into LVOT physiology. Standard 2D CNNs trained on single frames performed modestly and generalized poorly, suggesting that static morphology alone is insufficient for robust LVOT prediction. In contrast, EchoPrime, designed to capture high-resolution temporal dynamics, performed consistently well. Physiologically, this aligns with the dynamic nature of LVOTO, which reflects not only structural predisposition (septal thickness, leaflet length, LVOT diameter) but also systolic anterior motion, leaflet–septal interaction, and changes in LV cavity size throughout systole. Cross-attention fusion further improved performance by enabling the model to combine PLAX visualization of LVOT anatomy with apical-view information about flow direction and mitral valve kinematics. These findings reinforce that multi-view integration and temporal modeling are critical for reliable LVOT characterization.

The proposed multi-view LVOTO classification model has meaningful clinical utility. Our group has previously developed and validated an automated pipeline for detecting HCM from echocardiograms^17^. The proposed work extends this pipeline by providing a complementary tool to identify patients with obstructive HCM, specifically flagging patients with an LVOT gradient > 20mmHg, who would qualify for targeted treatment therapies. Integrating the gradient classification model with the existing HCM detection pipeline would allow automated identification of obstructive HCM, enabling clinicians to recognize a subgroup of patients who might otherwise be missed and ensure timely referral for appropriate management.

Furthermore, the proposed AI-guided strategy demonstrated substantial cost savings (3x) compared with a universal screening approach, primarily by reducing the number of patients requiring full echocardiogram along with provocation maneuvers where needed. This sequential framework preserves clinical performance while improving resource utilization. Notably, these estimates may be conservative, as real-world constraints such as limited access to specialized centers and associated delays were not incorporated into the analysis.

Additionally, this framework assumes that all false negative cases result in unexpected hospitalization, representing a worst-case scenario that likely overestimates the true downstream cost burden. Together, these models lay the groundwork for an end-to-end, fully automated echocardiographic workflow capable of screening, diagnosing, and phenotyping HCM patients at scale. An automated model capable of identifying elevated LVOT gradients from non-Doppler TTE could serve as a real-time decision-support tool during routine scanning. This approach may extend LVOT gradient assessment to settings where full Doppler evaluation is not feasible, including handheld or portable ultrasound devices. Furthermore, by generating probabilistic classifications across key clinical thresholds, the model represents a scalable approach for monitoring treatment efficacy. As shown in prior single-view work, DL-derived indices often follow the general direction of gradient changes after either medical or septal reduction therapy.

This study has several limitations. The datasets were retrospectively acquired from tertiary centers, and as such, additional validation in community settings, handheld imaging environments, and novice operators are required. Our model predicts gradients above or below 20 mmHg; future work should evaluate performance at traditional therapeutic thresholds (≥30 and ≥50 mmHg) and explore continuous gradient prediction. The study focuses on LVOTO in HCM; performance in non-HCM causes of dynamic obstruction or in patients with coexisting aortic stenosis remains unknown. Finally, while attention maps illustrate interpretable motion cues, further work is required to enhance model interpretability and clinician trust.

## Conclusion

This novel AI model with attention-based multi-view fusion applied to non-Doppler TTE videos can accurately classify LVOT gradient severity in HCM. The domain-specific Echo foundational model consistently outperformed conventional architectures and demonstrated strong external generalizability, while multi-view fusion further improved discrimination by leveraging complementary spatial–temporal information. These findings suggest that automated, video-based LVOT assessment may serve as a practical and scalable decision-support tool, complementing Doppler and expanding access to high-quality HCM evaluation.

## Data Availability

The data in this study can be made available upon reasonable request.

https://github.com/owencrystal1/GradientAI

## Abbreviations

AP3: Apical 3-Chamber
AP4: Apical 4-Chamber
AUROC: Area Under the Receiver Operating Curve
CNN: Convolutional Neural Network
DL: Deep Learning
HCM: Hypertrophic Cardiomyopathy
LVOT: Left Ventricular Outflow Tract
LVOTO: Left Ventricular Outflow Tract Obstruction
PLAX: Parasternal Long Axis
TTE: Transthoracic Echocardiography

---

**Supplementary Figure 1.**
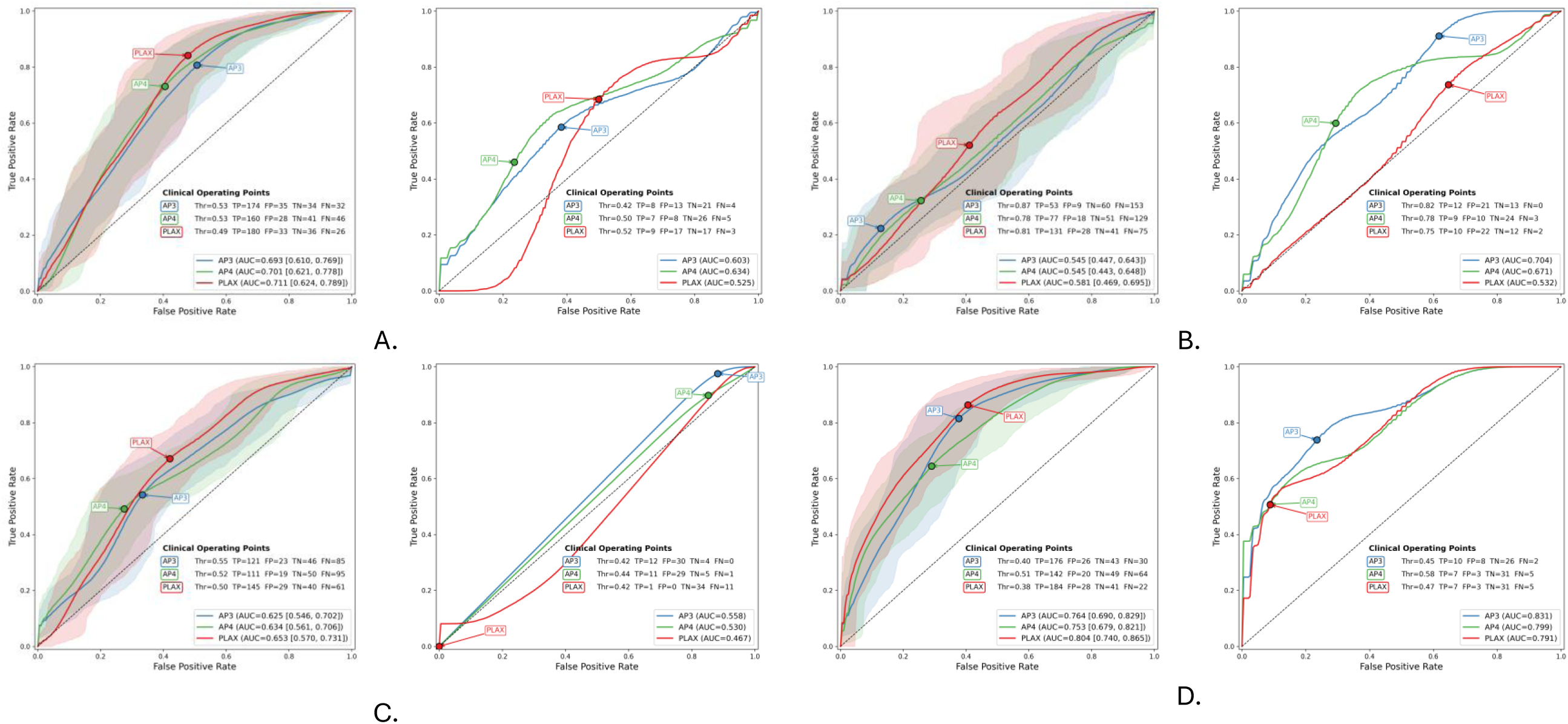
Quantitative performance in terms of AUROC on the internal (left) and external (right) datasets for the single-view 2D CNNs (A), single-view 3D CNNs (B), single-view 2D MedCLIP models (C), and single-view EchoPrime models (D).Optimal operating points and the corresponding number of true positives (TP) and false positives (FP) are listed for each model.

**Supplementary Figure 2.**
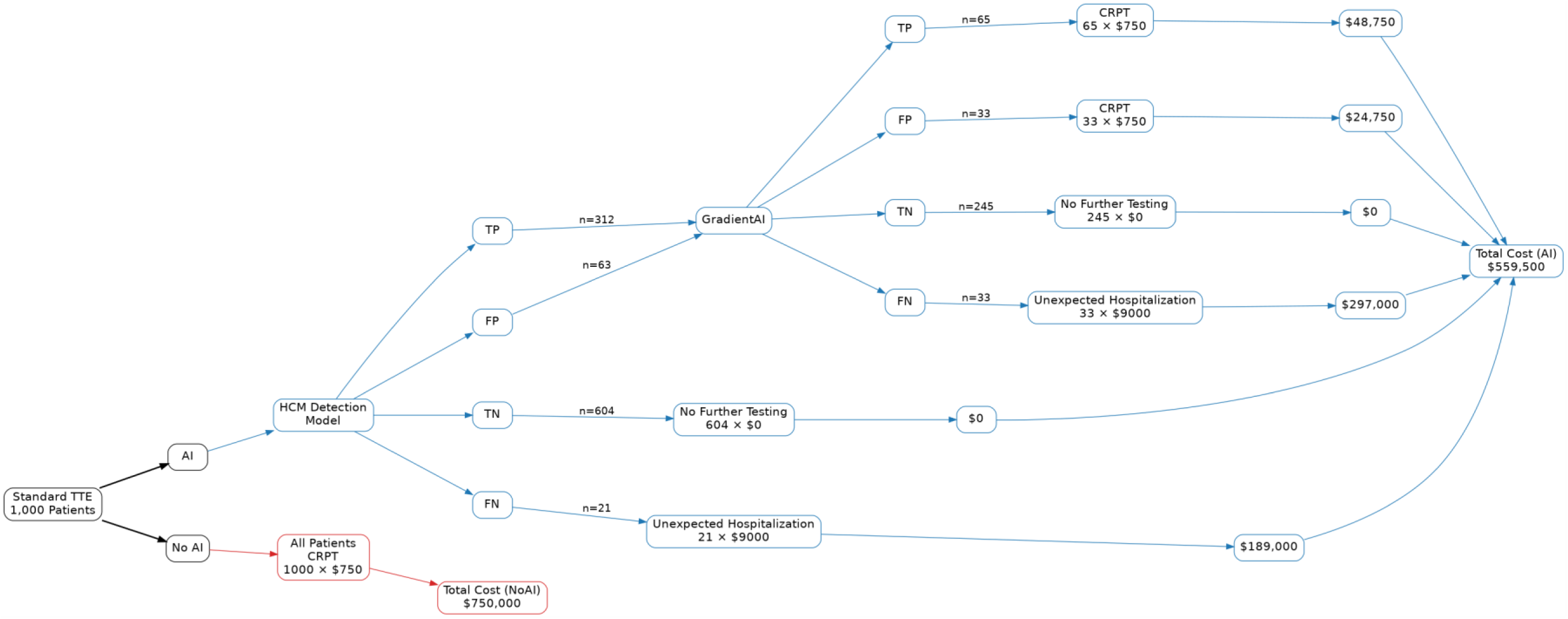
Decision tree analysis evaluating the financial impact of a two-stage AI-guided screening strategy using Medicare/Medicaid-based cost estimates. Outcomes were projected for a hypothetical cohort of 1,000 patients undergoing standard transthoracic echocardiography (TTE), based on model performance and prevalence from the external test set. In the AI-guided strategy, patients first underwent HCM detection, followed by LVOT gradient assessment among those classified as positive, and received a comprehensive rest and provocative TTE (CRPT). This sequential approach reduced downstream testing and resulted in substantially lower total costs compared with a “screen-all” strategy ($559,500 vs. $750,000), while maintaining clinical performance.

**Supplementary Figure 3.**
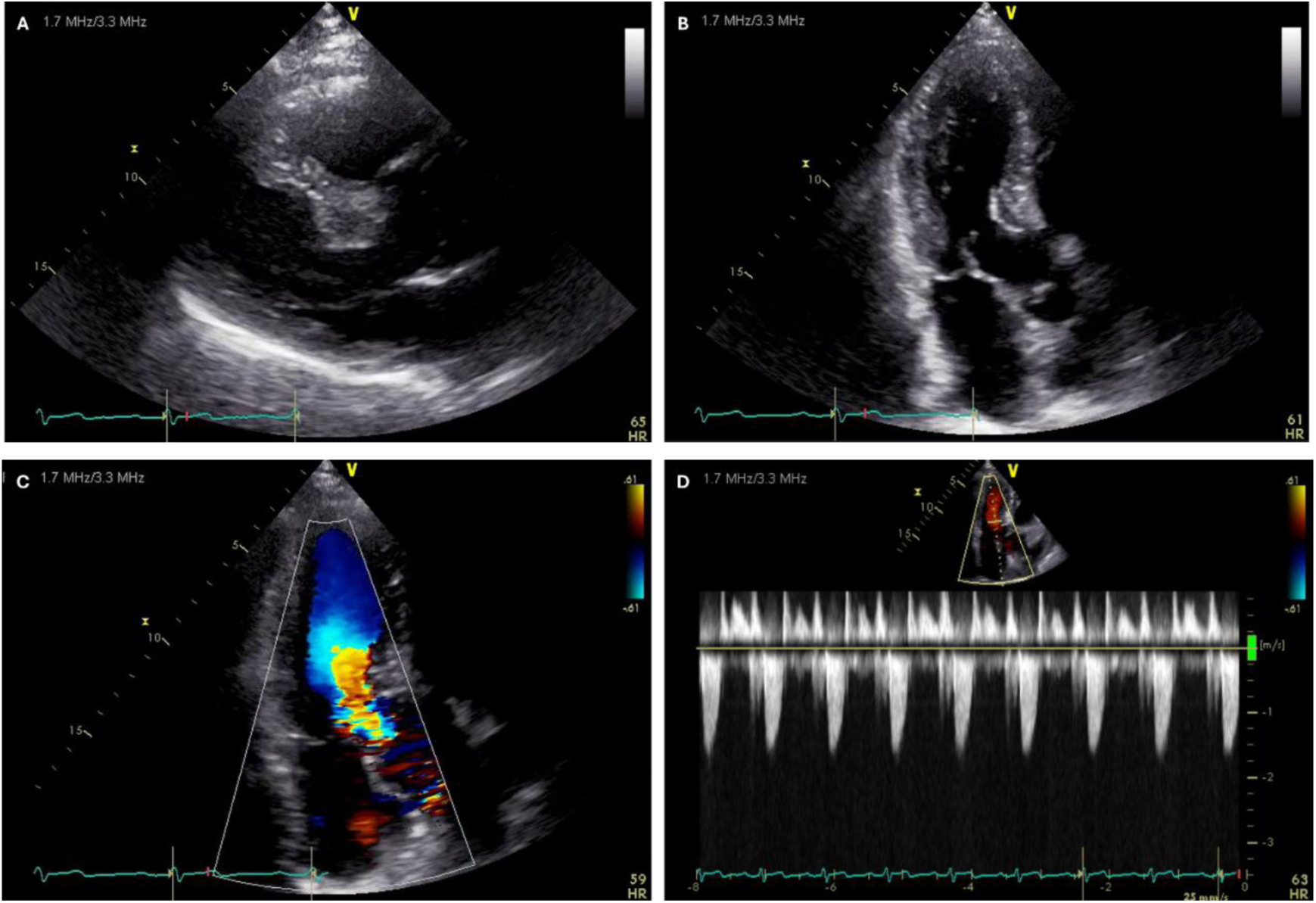
Example of a false-positive prediction Representative transthoracic echocardiographic images from a patient with hypertrophic cardiomyopathy in whom the AI model predicted the presence of an elevated left ventricular outflow tract (LVOT) gradient. Shown are 2D images of parasternal long-axis (PLAX, Panel A) and apical three-chamber (AP3, Panel B) views, with color Doppler demonstrating mild flow acceleration and aliasing in the LVOT on the AP3 view (Panel C). Continuous-wave Doppler interrogation of the LVOT demonstrates a peak velocity of approximately 1.6 m/s (Panel D), corresponding to a modest increase in LVOT gradient that does not meet the predefined threshold for classification of significant obstruction. This case illustrates a false-positive AI prediction driven by subtle hemodynamic features suggestive of early or borderline LVOT flow acceleration.

